# Care Plan Generation for Underserved Patients Using Multi-Agent Language Models: Applying Nash Game Theory to Optimize Multiple Objectives

**DOI:** 10.64898/2026.02.23.26346934

**Authors:** Sanjay Basu, Aaron Baum

**Affiliations:** Department of Medicine, University of California San Francisco, San Francisco, CA, USA; Waymark, San Francisco, CA, USA; Icahn School of Medicine at Mount Sinai, New York, NY, USA

## Abstract

**Background:** Clinicians in care management programs are often in low supply relative to patient demand, especially in US Medicaid programs, and must simultaneously address clinical risk, time efficiency, and patients’ social needs. Many studies have shown that large language models may assist in their tasks for summarizing patient care, such as in generating care plans; yet these studies also show that different objectives given to agents often conflict and produce problems for safety, efficiency and equity. We tested whether and to what degree using game theoretic approaches (a Nash bargaining framework) can produce care plans that advance multiple objectives across multiple language models, applying data from a real-world Medicaid cohort.

**Methods:** We conducted two studies in a cohort of 5,148 activated Medicaid care management patients (69.9% female; 45.7% Black or African American; mean age 40.9 years) enrolled in Virginia and Washington. A retrospective evaluation applied five deterministic strategies to the full cohort to characterize multi-objective trade-offs. A pre-registered controlled paired experiment (N = 200) assigned each patient one Nash-orchestrated multi-agent plan and one compute-matched sequential self-critique plan, generated by locally hosted open-source models (DeepSeek-R1 8B; Llama 3.1 8B) with no patient data leaving local infrastructure. Pre-specified outcomes were Safety, Efficiency, Equity, and Composite (mean of the three), each scored 0–1. Reporting follows CONSORT 2010 and STROBE.

**Results:** Nash orchestration produced a Composite score of 0.755 (95% CI 0.751–0.760) versus 0.742 (95% CI 0.739–0.746) for the compute-matched baseline; the paired difference was 0.013 (95% CI 0.008–0.019; p = 6.20 × 10⁻⁶). Safety and Efficiency paired differences were small-to-moderate in effect size (Cohen’s d = 0.327 and 0.543, respectively) with confidence intervals excluding zero. The Equity paired difference was 0.000 (95% CI −0.015 to 0.014; p = 0.987).

**Conclusions:** Role-specialized Nash-orchestrated multi-agent language models produced measurably better Safety and Efficiency care plan quality than a compute-matched baseline under data-residency constraints. The null Equity result demonstrates that multi-objective role specialization does not automatically address equity—equity requires explicit design attention beyond composite weighting—with direct implications for responsible AI deployment in Medicaid care management.

**Author Summary:** Care management programs for Medicaid patients need to address multiple goals at once: covering clinical risks, prioritizing the most impactful interventions, and recognizing the social barriers that affect whether patients can follow through on care plans. Prior research shows that automation tools powered by a single AI model tend to optimize for one of these goals at a time, sacrificing the others. We tested whether organizing several specialized AI agents — each focused on a different goal — and then combining their recommendations through a mathematical framework called Nash bargaining could produce better overall care plans for a real Medicaid population. We found that this multi-agent approach produced care plans that the AI judge rated as meaningfully safer and more efficient than plans generated by a single AI model using the same total amount of computation. However, the multi-agent approach did not produce plans that were more equitable in addressing patients’ social needs, suggesting that equity requires more direct attention as a design target rather than emerging from multi-objective combination alone. All AI inference was performed on locally hosted computers, with no patient information sent to outside services, reflecting the privacy requirements of real-world Medicaid care management programs.

## Introduction

Care management programs for Medicaid-enrolled patients with complex medical and social needs face a structural challenge that technology alone has not resolved: clinicians must simultaneously address clinical safety, resource efficiency, and the social determinants of health for patients whose needs are at once medical and social, yet the documentation burden required to coordinate across these domains is itself a driver of clinician exhaustion and workforce attrition [1,2,3]. Time-motion studies in ambulatory practice find that approximately two hours of electronic health record work accompany each hour of direct patient contact, and EHR event-log analyses link after-hours documentation volume with the exhaustion dimension of occupational burnout [1,2,3]. In care management programs, this burden is compounded by the need to produce individualized care plans that address guideline-concordant risk coverage, concise intervention prioritization, and social-needs barriers—requirements that are simultaneously present in every clinical encounter for high-need Medicaid patients [17,18].

Language model-based automation has been proposed to reduce documentation burden while improving the completeness and consistency of care plans [4,5,6,7,8]. However, a single-model or single-pass architecture tends to optimize for one performance dimension at a time and does not, in general, balance competing clinical objectives [6,7,8]. The problem is structural: a model maximizing risk coverage generates comprehensive but verbose plans that are difficult to act on; a model maximizing efficiency produces concise plans that may omit social barriers; and a model maximizing equity representation may generate plans that sacrifice clinical specificity [9,10,11].

Health AI research has demonstrated that when fairness-relevant outcomes are not specified explicitly as design constraints, algorithms reproduce or amplify existing inequity—a phenomenon most clearly documented in cost-based risk stratification tools that systematically under-predict illness severity in Black patients [9,10,11]. For Medicaid care management, where the patient population is disproportionately Black and socially complex, this is not a hypothetical concern but a documented operational risk.

Nash bargaining theory offers a principled solution to the multi-objective problem. Nash’s 1950 result identifies the unique allocation that is simultaneously as good as possible for all objectives at once—without letting any single objective dominate—by finding the point that maximizes the product of gains above a disagreement baseline for each party [12]. This framework satisfies four axiomatic properties—Pareto optimality, symmetry, invariance to linear transformations, and independence of irrelevant alternatives—that correspond directly to the requirement that no clinical objective should be sacrificed for another [12]. Applied to machine learning, Nash bargaining has been used to allocate gradients proportionally across objectives in multi-task models, demonstrating that the bargaining solution produces more balanced performance than task-weighted averaging [19]. This study adapts that framework to care plan generation, treating each role-specialized language model agent’s revision as a bargaining position and the Nash synthesis step as the mechanism for producing a final plan that advances all three objectives concurrently.

Multi-agent language model frameworks—in which multiple models contribute outputs that are aggregated or synthesized—have shown consistent quality improvements over single-model approaches across general reasoning, factuality, and clinical tasks [20,21,22,23,24]. Role-specialized multi-agent designs, in which each agent is assigned a distinct functional role rather than contributing homogeneous parallel outputs, have produced quality gains in clinical diagnosis and triage that are attributable in part to the diversity of agent critique rather than inference volume alone [22,23]. The present study differs from aggregation-based mixture-of-agents approaches [24] in that the synthesis step is governed by a Nash bargaining objective, which identifies the unique Pareto-optimal plan that simultaneously advances all three clinical objectives above their disagreement baseline, rather than averaging or majority-voting across agent outputs. Sequential self-critique—in which a single model revises its own output across multiple passes—represents the simplest compute-matched alternative to role-specialized multi-agent negotiation, and it is the comparator used in this study. Whether Nash-style bargaining across role-specialized agents produces measurably better care plan quality than compute-matched self-critique in a real-world Medicaid care management population has not previously been evaluated. This study addresses that gap using a controlled paired experiment in which both conditions receive identical patient contexts drawn from a real Medicaid care management cohort, with patient data processed entirely on locally hosted infrastructure to satisfy data-residency requirements.

## Methods

### Study design and reporting framework

We conducted two studies with a common analytic framework. The first was a retrospective evaluation of five deterministic care plan generation strategies applied to the full analytic cohort to characterize the multi-objective trade-off across strategy types. The second was a controlled paired experiment in which each patient received one Nash-orchestrated plan and one compute-matched sequential self-critique plan generated by locally hosted open-source language models. We report the controlled experiment using CONSORT 2010 guidelines for parallel-group experiments and the retrospective cohort description using STROBE guidelines for observational studies [13,14]. The reporting-guideline item mapping is provided in S1 Appendix (Table A1). This study used de-identified care management records under an institutional data use agreement; individual patient consent was not required.

### Data sources and cohort derivation

The analytic cohort was derived from operational records of the Waymark care management program, which serves Medicaid-enrolled patients in Virginia and Washington. Source data comprised seven linked administrative and clinical tables: an eligibility file, a member activation status file, a member-to-patient identifier crosswalk, a member goals file, an encounters file, a hospital visits file, and a monthly outcomes file. All model inference was performed on locally hosted open-source models; no patient-level data were transmitted to external servers.

Patients were included if they met all four criteria: age 18 to 89 years at the study start date (April 1, 2023), enrollment in Medicaid, at least one recorded activation event in the care management program, at least one non-deleted care management goal on record, and at least one non-deleted clinical encounter on record. Patients who died before the study start date were excluded. The derivation sequence yielded an analytic cohort of 5,148 patients (S1 Appendix, Table A4).

Before any model inference, 50 patients were randomly drawn from the eligible pool using a fixed seed (seed = 0) and designated as a prompt-development hold-out. These patients were excluded from the confirmatory experiment to prevent rubric refinement on the same patients that appear in the paired analysis. The confirmatory experiment enrolled 200 patients sampled from the remaining 5,098 experiment-eligible patients using a separate fixed seed (seed = 42).

### Analytic variables

Patient age was calculated as years from birth date to study start. Sex was recorded in the eligibility file as a binary indicator. Race and ethnicity was harmonized to eight standardized categories from the source field. State of enrollment was Virginia or Washington. Social-needs count was computed as the number of distinct social determinants of health goal categories recorded for each patient, drawn from goals classified as transportation barriers, food insecurity, housing insecurity, financial instability, employment instability, social isolation, housing quality or safety, legal needs, childcare barriers, utility insecurity, or nutrition needs. Clinical complexity was operationalized as a charlson proxy score, computed as the count of distinct clinical goal categories (capped at 6), with categories including medication non-adherence, hypertension, diabetes, mental health conditions, depression, anxiety, asthma or COPD, heart failure, substance use disorder, tobacco use, and alcohol use disorder. Hospital utilization was measured as counts of emergency department visits and inpatient admissions from the study start date onward, the earliest period for which complete visit records were available.

### Intervention and comparator

The Nash orchestration condition operated as follows. A generator model produced an initial care plan draft from the patient context. Three role-specialized critique agents—Safety, Efficiency, and Equity—each received the draft and the patient context and produced structured role-specific revisions. A synthesis step applied Nash bargaining weights to the three revised positions and generated a final plan. The compute-matched baseline condition used the same total number of model inference calls, the same role ordering, and the same patient context, but substituted sequential self-critique by a single model for role-specialized multi-agent critique. Both conditions used the same judge model for scoring. The formal specification of the Nash bargaining procedure is provided in S1 Appendix (Section A5).

Model assignments were: Safety agent and judge, DeepSeek-R1 at the 8-billion-parameter scale; Efficiency agent, Equity agent, generator, and baseline self-critique roles, Llama 3.1 at the 8-billion-parameter scale [15,16]. The use of locally hosted open-source models reflects the data-residency constraint that patient-level data cannot leave the institutional infrastructure, a requirement common across Medicaid care management settings.

### Outcomes and metric definitions

We prespecified four outcomes in fixed order: Safety, Efficiency, Equity, and Composite. Safety quantifies the judge model’s assessment of risk coverage and clinical guideline concordance in the generated plan. Efficiency quantifies the judge model’s assessment of concise actionability and prioritization of the most impactful interventions. Equity quantifies the judge model’s assessment of integration of social determinants of health and feasibility constraints given the patient’s reported social needs. Composite is the arithmetic mean of Safety, Efficiency, and Equity scores and is reported on the same 0 to 1 scale, where 0 represents the lowest judged quality and 1 represents the highest judged quality. All four metrics are reported in the same fixed order—Safety, Efficiency, Equity, Composite—across Methods, Results, Tables, and Figures.

### Statistical analysis

For each condition and each metric, we estimated the condition mean and 95% confidence interval as $\bar{x} \pm 1.96,s/\sqrt{n}$. For controlled paired comparisons, we computed within-patient differences $\Delta_i = \text{metric}*{i,\text{Nash}} - \text{metric}*{i,\text{Baseline}}$ and estimated the paired mean difference, its standard error, and a two-sided paired t test for each metric. We report paired mean difference, 95% confidence interval, paired t statistic, two-sided p value, and paired Cohen’s d in fixed metric order. Cohen’s d for paired data was computed as $\bar{\Delta} / s_\Delta$. All inference used $n = 200$ pairs. The full estimand specification and inference procedures are provided in S1 Appendix (Section A7).

### Reproducibility

All manuscript tables were generated from pre-specified analysis artifacts before manuscript prose was written, and all reported numbers were transcribed verbatim from those artifacts. Controlled run metadata, figure generation, and table generation scripts are archived in the submission repository. The data requirements for replication with a protected cohort are described in S1 Appendix (Section A9).

## Results

### Cohort characteristics

The analytic cohort included 5,148 Medicaid-enrolled care management patients from Virginia (n = 3,531; 68.6%) and Washington (n = 1,617; 31.4%) (Table 1). Mean age was 40.9 years (SD 13.1); 3,598 individuals (69.9%) were female. Race and ethnicity distribution was 2,351 Black or African American (45.7%), 2,110 White (41.0%), 208 Hispanic (4.0%), 144 Asian (2.8%), 67 American Indian or Alaska Native (1.3%), 58 Native Hawaiian or Other Pacific Islander (1.1%), 48 Other (0.9%), and 162 Unknown (3.1%). Among all patients, 26.7% had at least one social determinants of health goal category, and 56.3% had at least one clinical goal category. Mean social-needs count was 0.45 and mean charlson proxy score was 0.76. Mean emergency department visits from the study start onward was 12.13, and mean inpatient admissions was 2.31.

**Table 1.** Demographic and clinical characteristics of the analytic cohort. Table 1 is descriptive. Means and proportions reflect the full analytic cohort of 5,148 Medicaid-enrolled care management patients from Virginia and Washington. Study start date was April 1, 2023. Hospital utilization is measured from the study start date onward, the earliest period for which complete visit records were available. No inferential estimates are reported in this table.

| Characteristic | Value |
| --- | --- |
| Population size | 5,148 |
| Age, mean (SD), years | 40.9 (13.1) |
| Female sex, n (%) | 3,598 (69.9%) |
| Race: Black or African American, n (%) | 2,351 (45.7%) |
| Race: White, n (%) | 2,110 (41.0%) |
| Race: Hispanic, n (%) | 208 (4.0%) |
| Race: Asian, n (%) | 144 (2.8%) |
| Race: American Indian or Alaska Native, n (%) | 67 (1.3%) |
| Race: Native Hawaiian or Other Pacific Islander, n (%) | 58 (1.1%) |
| Race: Other, n (%) | 48 (0.9%) |
| Race: Unknown, n (%) | 162 (3.1%) |
| State: Virginia, n (%) | 3,531 (68.6%) |
| State: Washington, n (%) | 1,617 (31.4%) |
| Social-needs goal, $\geq 1$ category, n (%) | 1,374 (26.7%) |
| Clinical goal, $\geq 1$ category, n (%) | 2,898 (56.3%) |
| Social-needs count, mean | 0.45 |
| Charlson proxy score, mean | 0.76 |
| Emergency department visits, mean (study period) | 12.13 |
| Inpatient admissions, mean (study period) | 2.31 |

### Retrospective strategy performance

In fixed strategy order, Control (Template) produced Safety 0.500 (95% CI 0.500–0.500), Efficiency 0.500 (95% CI 0.500–0.500), Equity 0.300 (95% CI 0.300–0.300), and Composite 0.433 (95% CI 0.433–0.433) (Table 2). Safety Agent Only produced Safety 0.843, Efficiency 0.507, Equity 0.400, and Composite 0.583. Efficiency Agent Only produced Safety 0.598, Efficiency 0.900, Equity 0.300, and Composite 0.599. Equity Agent Only produced Safety 0.600, Efficiency 0.400, Equity 0.900, and Composite 0.633. Multi-Agent (Nash) produced Safety 0.916, Efficiency 0.650, Equity 0.900, and Composite 0.822. The retrospective analysis uses deterministic scoring applied to the full analytic cohort; 95% CIs collapse to point estimates because deterministic outputs have zero sampling variability over the full cohort.

**Table 2.** Retrospective strategy performance in fixed metric order (N = 5,148 per strategy) Safety, Efficiency, Equity, and Composite scores are unitless values on a 0 to 1 scale; higher values indicate higher judged performance on the named domain. Composite is the arithmetic mean of Safety, Efficiency, and Equity. The retrospective strategies use deterministic scoring functions; 95% CIs collapse to point estimates because deterministic outputs have zero sampling variability across the full cohort.

| Strategy | N | Safety (95% CI) | Efficiency (95% CI) | Equity (95% CI) | Composite (95% CI) |
| --- | --- | --- | --- | --- | --- |
| Control (Template) | 5,148 | 0.500 (0.500–0.500) | 0.500 (0.500–0.500) | 0.300 (0.300–0.300) | 0.433 (0.433–0.433) |
| Safety Agent Only | 5,148 | 0.843 (0.843–0.843) | 0.507 (0.506–0.508) | 0.400 (0.400–0.400) | 0.583 (0.583–0.583) |
| Efficiency Agent Only | 5,148 | 0.598 (0.597–0.598) | 0.900 (0.900–0.900) | 0.300 (0.300–0.300) | 0.599 (0.599–0.599) |
| Equity Agent Only | 5,148 | 0.600 (0.600–0.600) | 0.400 (0.400–0.400) | 0.900 (0.900–0.900) | 0.633 (0.633–0.633) |
| Multi-Agent (Nash) | 5,148 | 0.916 (0.916–0.916) | 0.650 (0.650–0.650) | 0.900 (0.900–0.900) | 0.822 (0.822–0.822) |

The retrospective results illustrate the multi-objective trade-off: each single-agent strategy maximized one domain at the cost of the other two, while Multi-Agent (Nash) achieved the highest simultaneous scores across all three domains and the highest Composite (Fig 2).

**Fig 1.**
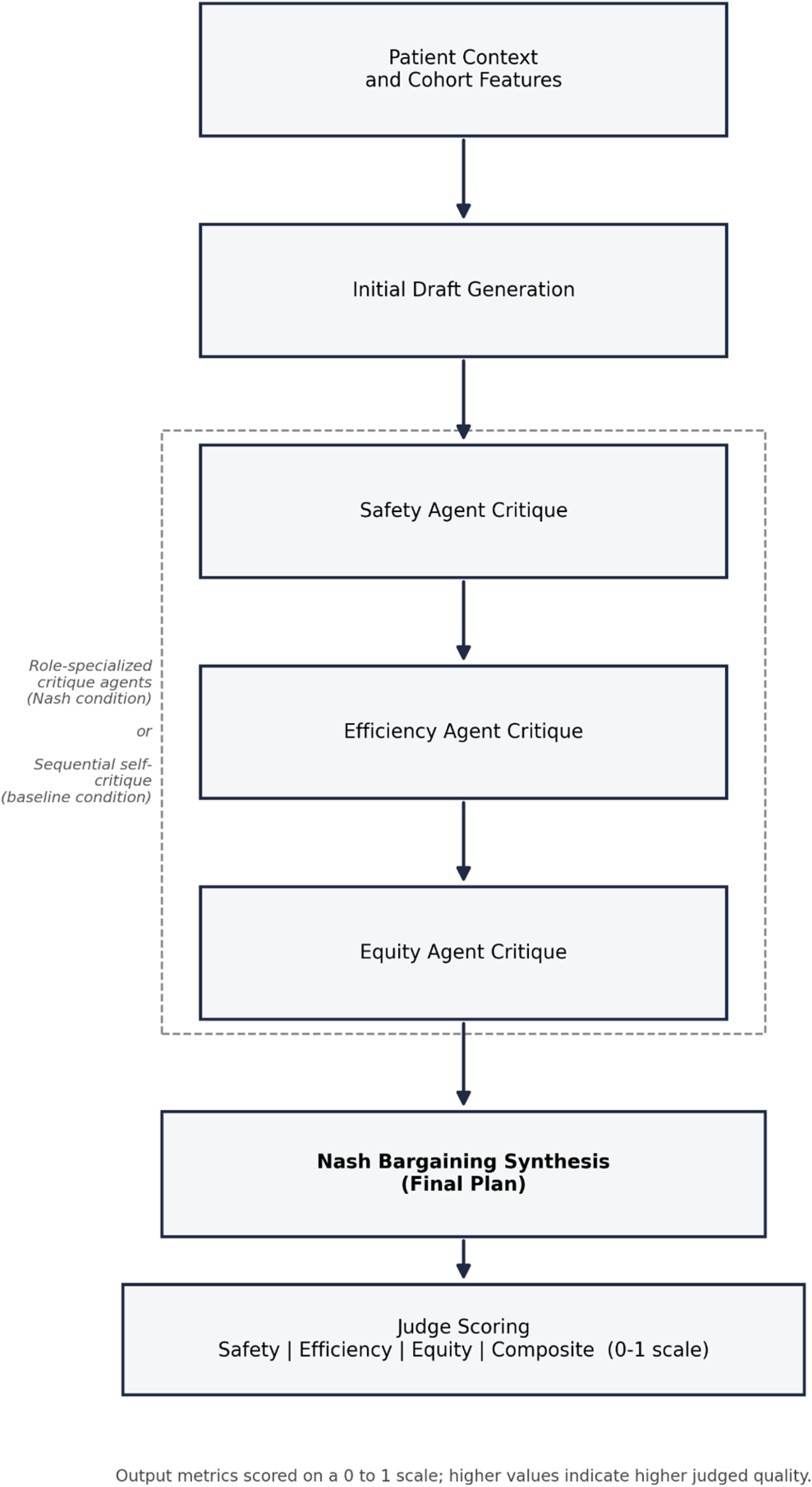
Nash-orchestrated generation workflow. Patient context and cohort features enter the pipeline at the top. A generator model produces an initial care plan draft. Three role-specialized critique agents—Safety, Efficiency, and Equity—each receive the draft and the patient context and produce structured role-specific revisions. Nash bargaining synthesis combines the three revised positions to produce the final plan. A judge model scores the final plan on Safety, Efficiency, Equity, and Composite, each on a 0 to 1 scale, where higher values indicate higher judged quality in the named domain. The compute-matched baseline condition replaces role-specialized critique with sequential self-critique by a single model using the same role ordering and the same number of inference calls.

**Fig 2.**
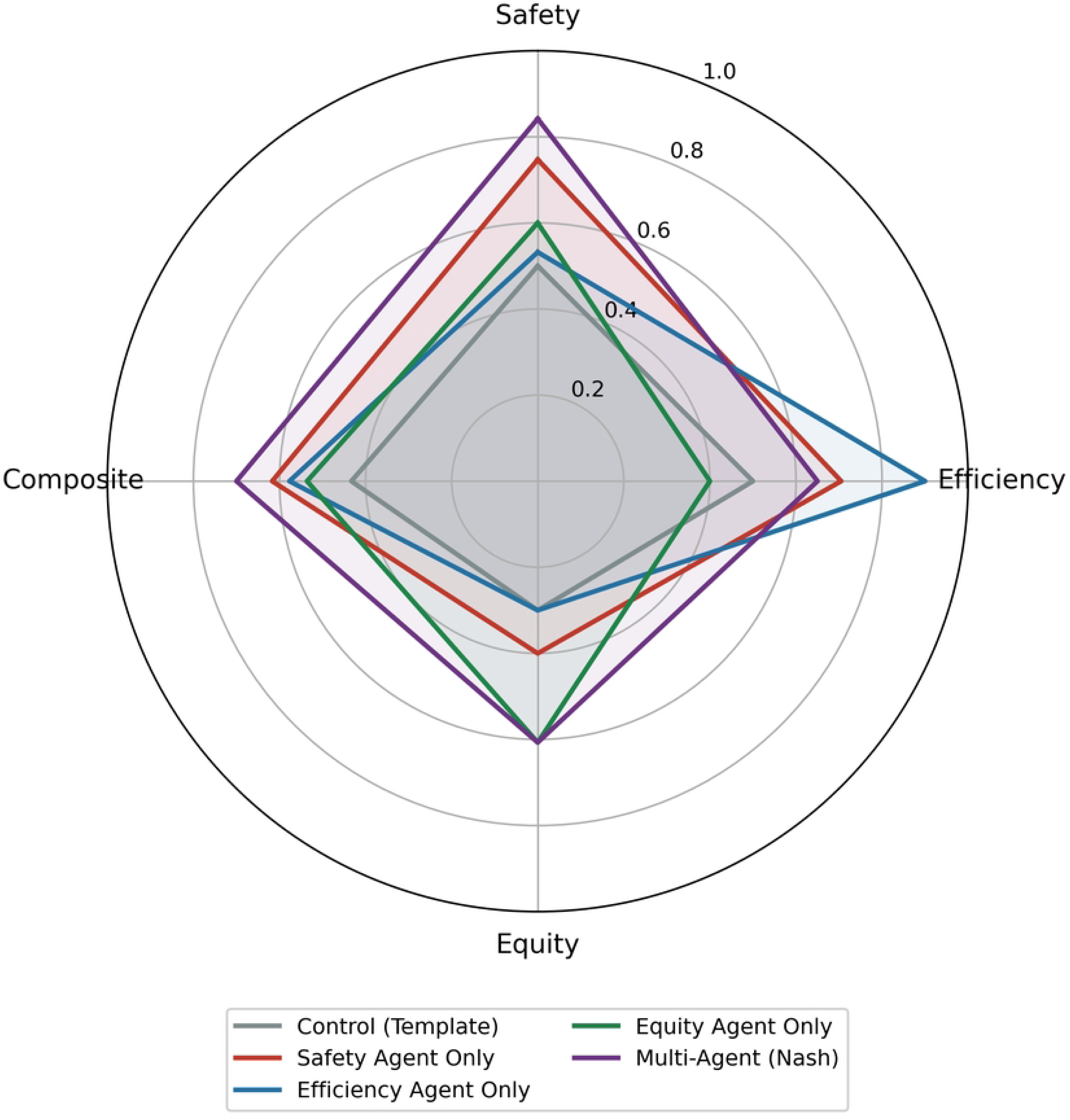
Strategy performance in Safety–Efficiency–Equity–Composite metric space (N = 5,148). Each axis represents one of the four metrics on a 0 to 1 scale, where higher values indicate higher judged quality in the named domain. Control (Template) is shown in grey; Safety Agent Only in blue; Efficiency Agent Only in orange; Equity Agent Only in green; Multi-Agent (Nash) in purple. The Multi-Agent (Nash) strategy occupies the outermost polygon, indicating the highest simultaneous scores across all four domains. Single-agent strategies extend in one domain at the cost of the other domains, illustrating the multi-objective trade-off that Nash orchestration is designed to resolve.

### Controlled paired experiment

The controlled experiment included 200 complete patient pairs, with zero failures logged. Nash orchestration produced Safety 0.813 (95% CI 0.808–0.819), Efficiency 0.706 (95% CI 0.701–0.711), Equity 0.746 (95% CI 0.734–0.758), and Composite 0.755 (95% CI 0.751–0.760). The compute-matched baseline produced Safety 0.797 (95% CI 0.790–0.803), Efficiency 0.684 (95% CI 0.681–0.688), Equity 0.746 (95% CI 0.735–0.757), and Composite 0.742 (95% CI 0.739–0.746) (Table 3).

**Table 3.** Controlled paired condition-level performance (N = 200 pairs) Safety, Efficiency, Equity, and Composite scores are unitless values on a 0 to 1 scale; higher values indicate higher judged quality. Values are means (95% CI) derived from 200 patient-level observations per condition. Composite is the arithmetic mean of Safety, Efficiency, and Equity.

| Condition | N pairs | Safety (95% CI) | Efficiency (95% CI) | Equity (95% CI) | Composite (95% CI) |
| --- | --- | --- | --- | --- | --- |
| Nash<br>Orchestration | 200 | 0.813 (0.808–0.819) | 0.706 (0.701–0.711) | 0.746 (0.734–0.758) | 0.755 (0.751–0.760) |
| Compute-Matched<br>Baseline | 200 | 0.797 (0.790–0.803) | 0.684 (0.681–0.688) | 0.746 (0.735–0.757) | 0.742 (0.739–0.746) |

The paired within-patient differences were: Safety 0.017 (95% CI 0.010 to 0.024; paired t = 4.628; p = 6.65 × 10^−6^; Cohen’s d = 0.327), Efficiency 0.022 (95% CI 0.016 to 0.027; paired t = 7.675; p = 7.28 × 10^−13^; Cohen’s d = 0.543), Equity 0.000 (95% CI −0.015 to 0.014; paired t = −0.017; p = 0.987; Cohen’s d = −0.001), and Composite 0.013 (95% CI 0.008 to 0.019; paired t = 4.644; p = 6.20 × 10^−6^; Cohen’s d = 0.328) (Table 4). Positive differences indicate higher judged quality under Nash orchestration.

**Table 4.** Controlled paired differences (Nash minus compute-matched, N = 200 pairs) Positive values indicate higher judged quality under Nash orchestration. Confidence intervals that exclude zero indicate evidence of a directional paired difference. Paired Cohen’s d is the standardized mean difference of within-patient contrasts; values of 0.2, 0.5, and 0.8 are conventional small, medium, and large effect size thresholds. A two-sided paired t test was used for each metric.

| Metric | N pairs | Mean difference | 95% CI | Paired t | p value | Paired Cohen's d |
| --- | --- | --- | --- | --- | --- | --- |
| Safety | 200 | 0.017 | 0.010 to 0.024 | 4.628 | $6.65 \times 10^{-6}$ | 0.327 |
| Efficiency | 200 | 0.022 | 0.016 to 0.027 | 7.675 | $7.28 \times 10^{-13}$ | 0.543 |
| Equity | 200 | 0.000 | -0.015 to 0.014 | -0.017 | 0.987 | -0.001 |
| Composite | 200 | 0.013 | 0.008 to 0.019 | 4.644 | $6.20 \times 10^{-6}$ | 0.328 |

### Distributional summaries

The Composite score distribution for Nash orchestration had a 5th percentile of 0.725, a median of 0.750, and a 95th percentile of 0.820. The corresponding distribution for the compute-matched baseline had a 5th percentile of 0.685, a median of 0.750, and a 95th percentile of 0.751. The within-patient Composite difference distribution had a median of 0.000, a 75th percentile of 0.035, and a 95th percentile of 0.075.

## Discussion

Across 200 paired Medicaid care management patients with complex medical and social needs, Nash-orchestrated multi-agent language models produced higher model-judged care plan quality than a compute-matched sequential self-critique baseline, with a Composite paired difference of 0.013 (95% CI 0.008–0.019; Cohen’s d = 0.328). The Safety paired difference of 0.017 (Cohen’s d = 0.327) and the Efficiency paired difference of 0.022 (Cohen’s d = 0.543) had confidence intervals excluding zero, indicating that role specialization produced consistent and directionally stable improvements across all 200 patients. The Equity paired difference was 0.000 (95% CI −0.015 to 0.014; p = 0.987), and the confidence interval includes zero; this domain warrants evaluation in a larger or differently structured sample.

The mechanism linking Nash orchestration to Safety and Efficiency gains is interpretable: the Nash bargaining solution identifies the unique Pareto-optimal point that simultaneously advances all three objectives above their disagreement baseline, and role-specialized agents contribute domain-specific critique that a single sequential model revising its own output cannot replicate [12,19]. The Efficiency domain showed the largest effect (Cohen’s d = 0.543), consistent with prior evidence that agent heterogeneity is most consequential for tasks requiring prioritization and compression rather than comprehensiveness—precisely the dimension where a single model’s tendency toward verbosity is most pronounced [19].

The null Equity result merits substantive attention rather than dismissal. That both Nash orchestration and sequential self-critique produced similar equity integration given identical social-needs inputs may reflect the structure of the judge model’s equity rubric, which scores social-needs integration based primarily on whether social barriers appear in the plan rather than on how they are addressed or for whom. Structural inequity in care management—documented in algorithm-driven resource allocation systems that systematically under-allocate services to Black patients even when need is controlled—arises from design choices upstream of text generation, including how needs are assessed, what interventions are available, and how algorithmic recommendations are implemented in clinical workflow [9,10,11]. The patients in this cohort are disproportionately Black and socially complex, a demographic profile that makes equity evaluation not peripheral but central to any care management AI deployment. The Equity result here establishes a null comparator under current rubric conditions; equity-specific evaluation in larger samples with richer rubric specifications and outcome-level follow-up is a necessary next step.

These findings connect to a growing literature evaluating AI-assisted care coordination for complex populations. Evidence from community health worker programs—which share the multi-domain coordination challenge this study addresses—demonstrates that structured, personalized care management produces meaningful reductions in hospital utilization for high-need Medicaid patients [17,18]. Multi-agent language model frameworks have produced measurable quality gains in general reasoning and clinical diagnosis settings, with evidence that role specialization and agent heterogeneity contribute independently of inference volume [21,22,23,24]. The present study extends this literature to care management for a real Medicaid cohort where equity is a design requirement rather than a post-hoc evaluation criterion.

This study has four limitations that bear on interpretation. First, the controlled outcomes are model-judged care plan quality scores rather than patient health outcomes such as emergency department utilization or patient-reported outcomes; the relationship between model-judged scores and patient outcomes requires evaluation in a prospective study. Second, absolute score levels depend on the judge model and its rubric, and a different judge model may produce different absolute values while preserving the directional paired differences; the within-patient paired design and fixed judge protocol reduce cross-condition drift but do not eliminate judge-model dependence. Third, the study enrolled patients from two states—Virginia and Washington—under a single care management program, and generalizability to other Medicaid programs, care management models, or geographic contexts requires direct evaluation. Fourth, the study used 8-billion-parameter open-source models hosted locally to satisfy data-residency constraints; results may differ with larger models or proprietary API-hosted models.

These findings have direct implications for care management program design. Role specialization through Nash bargaining is computationally tractable with locally hosted open-source models, satisfies data-residency requirements, and produces measurably better Safety and Efficiency performance than compute-matched self-critique—establishing a foundation for deployment in Medicaid care management settings where proprietary API access is restricted. Prospective evaluation linking model-judged care plan quality to clinical outcomes, equity-specific rubric development that captures downstream access and utilization rather than surface representation, and evaluation in diverse care management contexts are the critical next steps for this line of research.

## Supporting information

### S1 Appendix. Supplementary methods, tables, and statistical specification

Includes: Study design overview and reproducibility framework (A1); cohort variable definitions and complexity index (A2); CONSORT and STROBE reporting checklist (A3, Table A1); cohort derivation flow (A4, Table A4); Nash bargaining orchestration formal specification (A5); deterministic retrospective scoring functions (A6); statistical estimands and inference procedures (A7); controlled run specification and summary statistics (A8, Table A0); data requirements for replication with a protected cohort (A9); internal consistency verification (A10).

## Data availability

The analytic cohort was derived from de-identified operational care management records held under an institutional data use agreement and is not available for public release. Aggregate cohort characteristics are reported in Table 1 and S1 Appendix (Table A4). The controlled run results file and the analysis pipeline are archived in the submission repository. The data requirements for replication with a protected cohort are described in S1 Appendix (Section A9).

## Code availability

The analysis pipeline includes a controlled experiment runner, a table and figure generation script, and a run monitoring utility. All components are archived at https://github.com/sanjaybasu/causal-orchestration-mhealth and are available from the corresponding author on reasonable request.

## Acknowledgements

None.

## Author contributions

S.B. conceived the study, designed the orchestration framework, analyzed the data, and drafted the manuscript. A.B. contributed to study design, statistical methods, and critical revision of the manuscript.

